# Alzheimer^’^s-associated inflammatory alterations mediate tau-associated neurodegeneration in limbic and temporal regions across clinical variants of Alzheimer^’^s disease

**DOI:** 10.1101/2024.11.01.24316493

**Authors:** Patrick Lao, Seonjoo Lee, Daniel Talmasov, Dina Dass, Nbdusi Chikwem, Aubrey Johnson, Anna Smith, Diana Guzman, Amarachukwu Okafor, Hannah Houlihan, Lauren Heuer, Thairi Sanchez, Samantha Rossano, William Kreisl, James Noble, Yasir Qureshi, Scott Small

## Abstract

**Objectives:** Microglia monitor and respond to the brain^’^s microenvironment to maintain homeostasis. However, in Alzheimer^’^s disease and related dementias (ADRD), microglia may contribute to pathology. We hypothesized that AD-related inflammatory changes, measured with TSPO PET, would be locally associated with amyloid, tau, and neurodegeneration, and influence key pathways among them.

**Methods:** Participants (21 controls, 25 with ADRD) from the Longitudinal Imaging of Microglial Activation in Different Clinical Variants of Alzheimer^’^s Disease study underwent baseline amyloid PET (Florbetaben SUVR), tau PET (MK6240 SUVR), TSPO PET (ER176 SUVR), and structural MRI (gray matter volume). Cognitive assessments and consensus diagnoses (e.g., MCI, AD, PCA, FTD, LATE) were performed at the CUIMC ADRC with biomarker information when available. We evaluated regional colocalization of biomarker elevation in ADRD compared to controls, TSPO associations with ATN biomarkers, and TSPO mediations along key ATN pathways. Sensitivity analyses were stratified by amyloid positivity.

**Results:** Elevated TSPO was spatially colocalized with elevated tau (8 regions), amyloid (7 regions), and neurodegeneration (4 regions). Higher TSPO in limbic, temporal, and parietal regions was associated with higher tau (0.8 to 2.3, p<0.03), which remained significant after adjusting for amyloid and neurodegeneration in the inferior parietal cortex. TSPO mediated the association between tau and neurodegeneration in limbic and temporal regions (−0.27 to -0.39, ^p<0.02; 43%^ to ^89%^ of the total effect), while tau did not mediate the association between TSPO and neurodegeneration. TSPO also mediated the association between amyloid and tau, as well as tau across progressive Braak stages, but only in amyloid-positive ADRD.

**Conclusion:** Across ADRD diagnoses with different underlying brain microenvironments (e.g., pathology/copathology) to which microglia are sensitive, higher microglia density was associated with greater tau burden and mediated tau-associated neurodegeneration. Glia may represent a promising target for intervention strategies in ADRD-associated tau and neurodegeneration.

## Introduction

A comprehensive framework for Alzheimer^’^s disease (AD) that integrates proteopathic processes (such as the misfolding, aggregation, and spreading of amyloid and tau) with immunopathic mechanisms (including molecular and cellular components) could uncover alternative modifiable targets for preventing neurodegeneration and cognitive decline, beyond the traditional focus on amyloid and tau (for reviews, see [1, 2]). The initial case of AD noted glial cells with extensive fibrous structures and adipose saccules[3], later corroborated by genome-wide association studies implicating inflammation-related genes in AD and related dementias (ADRD)[4]. A chronic inflammatory environment is a hallmark of ADRD, leading to significant alterations in brain structure and function.

Microglia are one of the brain^’^s resident immune cells, continuously surveilling and responding to stimuli, including infection, toxins, and injury, thereby contributing to neuroprotection and repair. However, microglial function depends on the brain^’^s microenvironment, which can vary significantly under different physiological conditions. Studies employing TSPO PET imaging to assess microglial activity have revealed consistent increases in key brain regions across various neurodegenerative disorders, indicating potential disease-promoting mechanisms associated with microglia; however reductions in microglial density in key brain regions for autism spectrum disorder and various neuropsychiatric syndromes, suggests a lack of homeostatic maintenance in the absence of microglia[5]. Understanding both the disease stage (such as the extent of neurodegeneration and cognitive impairment) and the brain microenvironment (including amyloid and tau positivity and regional susceptibility) is essential for developing a unified framework that encompasses both proteopathic and immunopathic aspects of ADRD.

We utilize TSPO PET imaging to broadly capture ^“^AD-associated inflammatory alterations^”^ given that the 18kDa translocator protein (TSPO) is primarily expressed in microglia, as well as in astrocytes and infiltrating macrophages although at a lower level, and even in non-immune cell types such as endothelial cells[6-8]. TSPO PET may reflect microglial density and recruitment rather than function in humans[9]. This imaging technique has been shown to correlate with other inflammatory markers, including sTREM2[10]. The novel TSPO PET radiotracer ER176 exhibits favorable pharmacokinetic properties, including a specific non-displaceable binding ratio and a lack of radiometabolites entering the brain, which is an improvement over previous TSPO radiotracers[11, 12]. Notably, ER176 is less sensitive to the single nucleotide polymorphism in the TSPO gene (rs6971) that previously limited the signal to noise ratio of earlier TSPO PET radiotracers for approximately ^10%^ of the population[13]. Using semi-quantitative standard uptake value ratio (SUVR) with the cerebellum as a pseudo-reference region, we observed regional increases in TSPO for individuals with mild cognitive impairment (MCI) and AD compared to controls, aligning with gold standard distribution volume (VT) quantification obtained through arterial line sampling[14]. Other groups have shown the cerebellum to be a suitable pseudo-reference region on the pathological level[15], adding validation for its use with previous generation TSPO radiotracers[16]. Furthermore, the simplified PET acquisition method using ER176 is more amenable to older populations with ADRD.

Cross-sectionally, in MCI/AD, a previous TSPO PET radiotracer, PBR28, was associated with amyloid and tau burden, independently of neurodegeneration. It mediated the association between tau spreading across Braak stage regions, tau and neurodegeneration, and neurodegeneration and cognition[17]. Longitudinally, the pathways of tau propagation in MCI/AD depended on baseline PBR28 network strength, highlighting the role of microglia in tau propagation[10]. A recent study using ER176 demonstrated that TSPO PET was elevated in early onset MCI, a group characterized by a lack of expected copathology, compared to controls. TSPO PET exhibited a regional distribution similar to that of tau and neurodegeneration, with the strongest association found with tau and a correlation to memory impairment[18].

Here, we sought to generalize microglia-tau associations across various ADRD diagnoses that represent different brain microenvironments using simplified SUVR quantification for novel TSPO radiotracer, ER176. We hypothesized that within a cohort characterized at the Columbia Alzheimer^’^s Disease Research Center (ADRC) comprising individuals with MCI, AD, and posterior cortical atrophy (PCA) among other clinical variants, ER176 SUVR would be (1) elevated in ADRD across key brain regions similar to tau and neurodegeneration, (2) most strongly associated with tau, and (3) a mediator of progressive associations between amyloid, tau, and neurodegeneration.

## Methods

### Participants

Participants in the Alzheimer^’^s Disease Variant Imaging study (R01AG063888) were co-enrolled in the Columbia Alzheimer^’^s Disease Research Center (P30AG066462). Participants underwent amyloid PET, tau PET, TSPO PET, structural MRI, and ADRC cognitive testing. Briefly, participants were evaluated with the Mini-Mental State Examination (MMSE)[19], and domain-specific tests including the Selective Reminding Test[20], Trail Making Test[21], and categorical fluency[22]. Domain-specific cognitive test scores were transformed into z-scores using age-, sex-, and education-adjusted normative data derived for the National Alzheimer^’^s Coordinating Center Uniform Dataset (NACC)[23]. At CUIMC ADRC case consensus, participants were categorized as cognitively unimpaired (n=21), amnestic or non-amnestic mild cognitive impairment (MCI; n=7), AD dementia (n=7), PCA (n=5), lvPPA (n=1), LATE (n=1), and FTD (n=1). Three individuals with cognitive impairment have not yet gone to case consensus for a specific diagnosis, but scored 1.5 standard deviations below the normative sample and are included in the ADRD group. Biomarker data was used in case consensus when available. All participants (or their legally authorized representatives) provided informed consent according to the Declaration of Helsinki. The Institutional Review Board of Columbia University Irving Medical Center gave ethical approval for this work.

TSPO binding affinity was determined as previously described[24]. Briefly, genomic DNA from each subject was used to genotype the rs6971 polymorphism using a TaqMan assay[25]. Participants were high affinity binders (HAB; 55^%^), mixed affinity binders (MAB; 32^%^), or low affinity binders (LAB; 13^%^), aligning with previously reported frequencies[25]. APOE Genotyping was determined using the KASPar^®^ PCR SNP genotyping system (LGC Genomics) for the single nucleotide polymorphisms (SNPs) rs7412 and rs429358. Genotype data for these two SNPs were used to unambiguously define ^ε^2, ^ε^3, and ^ε^4 alleles. APOE information was only available in a subset of 17 participants (10 controls, 7 patients).

### Neuroimaging

Structural T1 MRI was performed in a 3T GE Signa Premier scanner (repetition time (TR): 6.6 ms, echo time (TE): 3 ms, voxel size = 1×1×1 mm^3^). Regions of interest were defined using the Hammers-N30R83-1 MM atlas in the PNEURO module of PMOD 3.9 (PMOD Technologies, [26]), except for the entorhinal cortex, which was defined using the Desikan-Killiany atlas in FreeSurfer 6.0 (Massachusetts General Hospital, Harvard Medical School; http://surfer.nmr.mgh.harvard.edu). Gray matter volumes were calculated for 13 AD-related regions (Prefrontal Cortex, Insula, Cingulate Gyrus, Fusiform, Lingual Gyrus, Entorhinal cortex, Middle Inferior Temporal Gyrus, Superior Temporal Gyrus, Inferior Parietal Cortex, Superior Parietal cortex, Amygdala, Hippocampus, and Striatum).

All PET scans were performed in Siemens Biograph64 mCT/PET scanner, reconstructed with OSEM (voxel size = 1×1×2 mm), and corrected for radioactive decay, attenuation, scatter, random events, and scanner deadtime and normalization. All preprocessing steps (i.e., frame realignment, coregistration with segmented MRI, SUVR calculation) and partial volume correction using the voxelwise Geometric Transfer Method (GTM) were performed in PMOD[27]. Amyloid PET was performed using Florbetaben (8.1 mCi; FBB) and FBB SUVR was calculated using 90-110 min data and cerebellar gray matter as reference region. Tau PET was performed using MK-6240 (5 mCi) and MK-6240 SUVR was calculated using 90-110 min data and inferior cerebellar gray matter as reference region. TSPO PET was performed using ER176 (up to 20 mCi) and ER176 SUVR was calculated using 60-90 min data and cerebellar gray matter as pseudo-reference region.

### Statistical analysis

Demographic characteristics, biomarker levels, and clinical characteristics were compared between controls and individuals with ADRD. For descriptive purposes, amyloid positivity was rated visually according to vendor instructions[28]. Tau positivity across Braak I/II (Hippocampus, Parahippocampal Gyrus) was categorized as 2 standard deviations above the mean of MK6240 SUVR in controls. Given the large range of ER176 SUVR in controls, TSPO positivity across a composite brain ROI were calculated as above the mean in controls by binding affinity groups (Supplemental Figure 1).

For analytic purposes, all biomarkers were used continuously. To assess regional biomarker elevations, we compared participants with ADRD to controls in separate ANCOVAs. To assess pairwise biomarker associations across ROIs, we included biomarker by ROI interactions in general linear models. To assess adjusted biomarker associations, we included amyloid, tau, and neurodegeneration by ROI interactions as simultaneous predictors of TSPO in general linear models. To assess key mechanistic pathways, we included TSPO mediations from amyloid to tau within ROIs (i.e., TSPO mediates amyloid-related tau); tau in earlier Braak stages to tau in subsequent Braak stages (i.e., TSPO mediates tau-related spreading); and tau to neurodegeneration within ROIs (i.e., TSPO mediates tau-related neurodegeneration). To support causal inference from cross-sectional mediations, opposite directions were also assessed (TSPO to amyloid to tau; TSPO to earlier Braak tau to later Braak tau; TSPO to tau to neurodegeneration). We performed a series of sensitivity models, stratifying by amyloid positivity, to understand the different brain microenvironments in which associations were present. All models were adjusted for age, sex, and body mass index (BMI)[29]. Models with ER176 SUVR were further adjusted for TSPO binding affinity and models with gray matter volume were further adjusted for intracranial total volume (ICV). All estimates are standardized for a measure of effect size and to facilitate comparison across models. Statistical models were adjusted for multiple comparisons using the multivariate t distribution or the False Detection Rate method, depending on the model, and run in R 4.4.1.

## Results

Controls and individuals with ADRD were similar in age, sex, BMI, race, ethnicity, TSPO affinity, and APOE Genotype (Table 1). As expected, individuals with ADRD had lower MMSE and cognitive domain scores. Out of 21 individuals without cognitive impairment 50% were amyloid negative, tau negative, TSPO negative and 50% were amyloid negative, tau negative, TSPO positive by definition. Out of 25 individuals with ADRD, 47% were amyloid positive, tau positive, and TSPO positive; 12% were amyloid positive, tau positive, and TSPO negative; 12% were amyloid negative, tau positive, and TSPO positive; and 29% were amyloid negative, tau negative, and TSPO positive (For biomarker positivity profile by ADRD diagnosis, see Supplemental Figure 2).

**Table 1.** Demographic characteristics for controls and individuals with ADRD.

|  | Control<br>N = 21 | ADRD<br>N = 25 | Test statistic,<br>P-value |
| --- | --- | --- | --- |
| Age | 70 (6) | 71 (8) | T=0.06,<br>p=0.96 |
| Sex |  |  |  |
| Women | 10 (48%) | 11 (44%) | $\chi^2<0.001$,<br>p>0.99 |
| Men | 11 (52%) | 14 (56%) |  |
| BMI | 25.1 (3.1) | 26.2 (5.3) | T=0.8,<br>p=0.42 |
| Education | 16.33 (2.08) | 16.44 (3.38) | T=0.13,<br>p=0.90 |
| Race |  |  |  |
| Asian | 0 (0%) | 1 (4.0%) | $\chi^2=3.6$<br>p=0.31 |
| Black | 3 (14%) | 1 (4.0%) |  |
| Mixed | 1 (4.8%) | 0 (0%) |  |
| White | 17 (81%) | 23 (92%) |  |
| Ethnicity |  |  |  |
| Hispanic | 2 (10%) | 2 (8.0%) | $\chi^2<0.001$,<br>p>0.99 |
| Non-Hispanic | 19 (90%) | 23 (92%) |  |
| MMSE | 29.0 (1.7) | 24.4 (3.9) | T=-5.0,<br>p=<0.001 |
| Episodic Memory | 0.26 (0.70) | -1.98 (0.86) | T=-9.2,<br>p<0.001 |
| Working Memory | -0.02 (0.81) | -1.45 (1.15) | T=-4.6,<br>p<0.001 |
| Fluency | 0.50 (0.58) | -1.29 (1.16) | T=-6.4,<br>p<0.001 |
| Visuospatial Ability | -0.23 (0.66) | -2.97 (2.63) | T=-4.7,<br>p<0.001 |
| TSPO Affinity |  |  |  |
| Low affinity | 12 (57%) | 13 (52%) | $\chi^2=0.43$<br>p=0.81 |
| Mixed | 7 (33%) | 8 (32%) |  |
| High affinity | 2 (9.5%) | 4 (16%) |  |
| APOE Genotype |  |  |  |
| E3/E3 | 12 (71%) | 2 (50%) | $\chi^2=4.5$<br>p=0.11 |
| E3/E4 | 5 (29%) | 1 (25%) |  |
| E4/E2 | 0 (0%) | 1 (25%) |  |
| Missing | 4 | 21 |  |

The spatial colocalization of elevated TSPO with elevated tau (8 regions) was greater than that with elevated amyloid (7 regions) and elevated neurodegeneration (4 regions; Figure 1). A unique region for elevated TSPO and tau colocalization was the amygdala, while the middle inferior temporal gyrus, prefrontal cortex, and inferior and superior parietal cortex also had elevated neurodegeneration and amyloid. The hippocampus only had elevated TSPO, striatum and insula only had elevated amyloid, and entorhinal cortex only had elevated tau.

**Figure 1.**
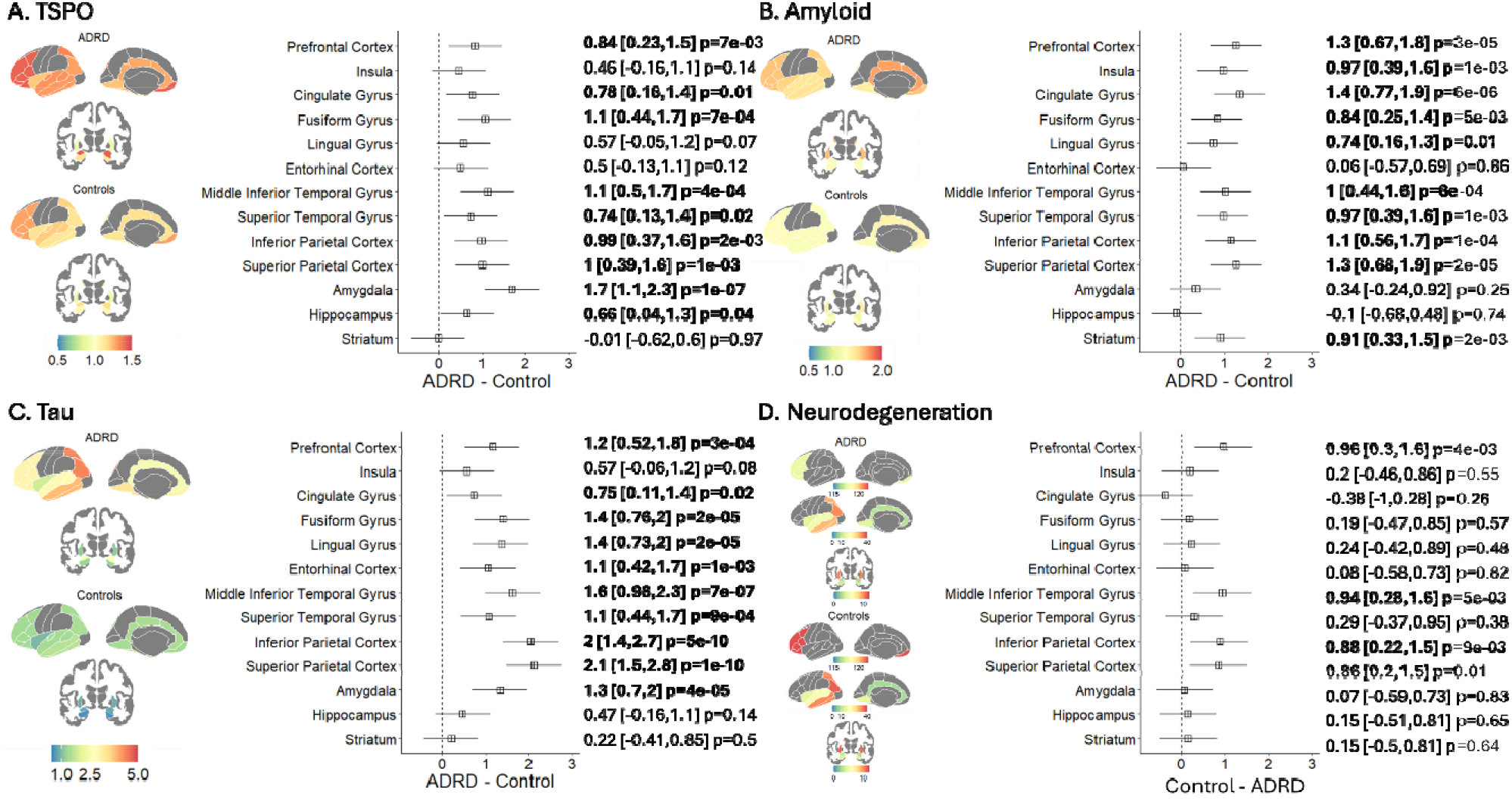
Average biomarker values for controls and individuals with ADRD as well as the difference between groups for (A) TSPO (ER176), (B) Amyloid (FBB), (C) Tau (MK6240), and (D) Neurodegeneration (Gray Matter Volume) across ROIs. Note that Neurodegeneration is displayed differently to accommodate a wide range of regional volumes and inverted (Control-ADRD) relative to other biomarkers for direct comparison across standardized effect sizes.

Further, greater TSPO in limbic, temporal, and parietal regions was associated with greater tau and with greater neurodegeneration (i.e., lower gray matter volume) in pairwise biomarker models (Figure 2). After adjustment for amyloid and neurodegeneration, greater TSPO was associated with greater tau in the inferior parietal cortex (0.3 [0.1, 0.5], p=0.05). After adjustment for amyloid and tau, greater TSPO was associated with greater neurodegeneration in the amygdala (−3.4 [-4.4, -2.4], p=2e-10), hippocampus (−1.9 [-2.6, -1.3], p=6e-08), and fusiform gyrus (−0.68 [-1.1, -0.28], p=0.01). Greater TSPO was associated with lower amyloid in the hippocampus, which did not survive adjustment for tau and neurodegeneration.

**Figure 2.**
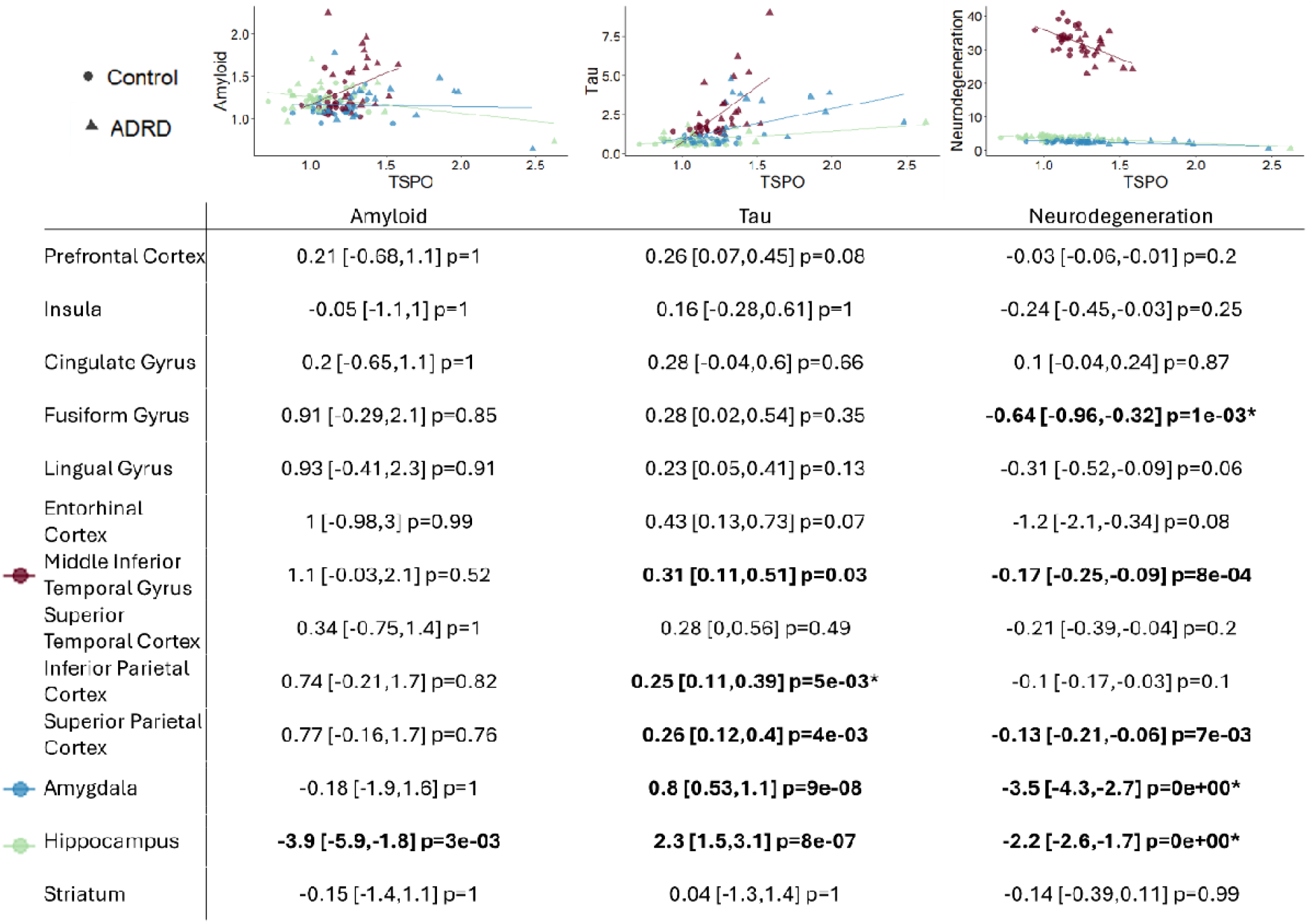
Scatterplots and pairwise biomarker models among TSPO and (A) Amyloid (FBB), (B) Tau (MK6240), and (C) Neurodegeneration (Gray Matter Volume) across ROIs. Circles represent Controls, Triangles represent ADRD, and colors represent select Regions of Interest (ROI). Asterisks indicate associations that survive adjustment in the multiple biomarker model

Assessing the role of TSPO in ATN progression, we found that TSPO mediated the association between tau and neurodegeneration in the middle inferior temporal gyrus (70% of total effect), amygdala (89%), and hippocampus (43%; Table 2), while the opposite mediation (tau mediating TSPO-associated neurodegeneration) was not present. However, in the cingulate gyrus, higher tau was associated with greater TSPO, which was associated with larger gray matter volume (despite a direct effect of greater tau with lower gray matter volume).

**Table 2.** TSPO mediation standardized estimates from a model of amyloid-associated tau (top), tau spreading (middle), and tau-associated neurodegeneration (bottom). Mediations from TSPO to Tau to Neurodegeneration were not significant.

|  | X->M | M->Y | X->Y | X->M->Y |
| --- | --- | --- | --- | --- |
|  | Amyloid->TSPO | TSPO->Tau | Amyloid-> Tau | Amyloid->TSPO->Tau |
| Prefrontal Cortex | 0.26 [-0.01,0.54] p=0.09 | **0.34 [0.11,0.57] p=7e-03** | **0.44 [0.22,0.67] p=2e-04** | 0.09 [-0.02,0.2] p=0.19 |
| Insula | 0.03 [-0.29,0.35] p=0.86 | 0.17 [-0.11,0.44] p=0.26 | **0.44 [0.17,0.7] p=1e-03** | 0 [-0.05,0.06] p=0.86 |
| Cingulate Gyrus | 0.29 [0.01,0.58] p=0.07 | 0.2 [-0.03,0.42] p=0.1 | **0.45 [0.22,0.67] p=2e-04** | 0.06 [-0.03,0.14] p=0.27 |
| Fusiform Gyrus | 0.05 [-0.27,0.38] p=0.81 | **0.21 [0.02,0.41] p=0.04** | **0.43 [0.24,0.62] p=3e-05** | 0.01 [-0.06,0.08] p=0.81 |
| Lingual Gyrus | **0.39 [0.09,0.68] p=0.04** | **0.29 [0.13,0.46] p=2e-03** | **0.44 [0.28,0.6] p=3e-07** | 0.11 [0.01,0.22] p=0.13 |
| Entorhinal Cortex | 0.29 [0.02,0.56] p=0.07 | **0.38 [0.1,0.65] p=0.01** | **0.5 [0.2,0.81] p=1e-03** | 0.11 [-0.02,0.24] p=0.19 |
| Middle Inferior Temporal Gyrus | **0.38 [0.07,0.69] p=0.05** | **0.32 [0.14,0.5] p=2e-03** | **0.36 [0.18,0.54] p=2e-04** | 0.12 [0,0.24] p=0.13 |
| Superior Temporal Cortex | 0.31 [0.02,0.6] p=0.07 | **0.27 [0.08,0.46] p=9e-03** | **0.41 [0.23,0.6] p=3e-05** | 0.08 [-0.01,0.18] p=0.19 |
| Inferior Parietal Cortex | **0.44 [0.18,0.7] p=0.01** | **0.34 [0.18,0.51] p=4e-04** | **0.49 [0.33,0.65] p=9e-09** | 0.15 [0.04,0.27] p=0.13 |
| Superior Parietal Cortex | **0.36 [0.1,0.63] p=0.03** | **0.28 [0.13,0.44] p=2e-03** | **0.52 [0.37,0.67] p=9e-11** | 0.1 [0.01,0.2] p=0.13 |
| Amygdala | -0.17 [-0.49,0.16] p=0.41 | **0.44 [0.22,0.65] p=4e-04** | **0.37 [0.16,0.57] p=6e-04** | -0.07 [-0.22,0.07] p=0.43 |
| Hippocampus | **-0.51 [-0.83,-0.19] p=0.01** | **0.37 [0.14,0.59] p=3e-03** | 0.05 [-0.18,0.28] p=0.68 | -0.19 [-0.35,-0.02] p=0.13 |
| Striatum | -0.08 [-0.37,0.21] p=0.7 | 0.08 [-0.22,0.38] p=0.6 | **0.5 [0.23,0.77] p=4e-04** | -0.01 [-0.04,0.03] p=0.81 |
|  | Tau->TSPO | TSPO->Tau | Tau->Tau | Tau->TSPO->Tau |
| Early and Middle Braak | **0.5 [0.19,0.8] p=1e-03** | **0.15 [0.03,0.28] p=0.03** | **0.75 [0.61,0.89] p=0e+00** | 0.08 [0,0.15] p=0.09 |
| Middle and Late Braak | **0.67 [0.38,0.97] p=1e-05** | 0.07 [-0.01,0.15] p=0.07 | **1 [0.91,1.1] p=0e+00** | 0.05 [-0.01,0.1] p=0.09 |
|  | Tau->TSPO | TSPO->Neurodegeneration | Tau->Neurodegeneration | Tau->TSPO->Neurodegeneration |
| Prefrontal Cortex | **0.52 [0.29,0.75] p=1e-05** | -0.13 [-0.46,0.19] p=0.42 | -0.26 [-0.58,0.06] p=0.11 | -0.07 [-0.24,0.1] p=0.43 |
| Insula | 0.16 [-0.17,0.49] p=0.35 | **-0.23 [-0.46,0] p=0.05** | **-0.29 [-0.52,-0.05] p=0.02** | -0.04 [-0.12,0.05] p=0.39 |
| Cingulate Gyrus | **0.48 [0.18,0.78] p=1e-03** | **0.53 [0.16,0.91] p=5e-03** | **-0.41 [-0.82,0] p=0.05** | **0.26 [0.02,0.5] p=0.04** |
| Fusiform Gyrus | **0.43 [0.04,0.81] p=0.03** | **-0.47 [-0.72,-0.21] p=4e-04** | -0.2 [-0.53,0.13] p=0.24 | -0.2 [-0.41,0.01] p=0.06 |
| Lingual Gyrus | **0.82 [0.5,1.1] p=4e-07** | -0.24 [-0.5,0.02] p=0.07 | -0.28 [-0.6,0.04] p=0.09 | -0.2 [-0.43,0.03] p=0.09 |
| Entorhinal Cortex | **0.33 [0.1,0.56] p=5e-03** | -0.04 [-0.32,0.23] p=0.76 | **-0.43 [-0.7,-0.16] p=2e-03** | -0.01 [-0.11,0.08] p=0.76 |
| Middle Inferior Temporal Gyrus | **0.78 [0.43,1.1] p=1e-05** | **-0.49 [-0.73,-0.26] p=4e-05** | -0.17 [-0.48,0.14] p=0.28 | **-0.39 [-0.64,-0.13] p=3e-03** |
| Superior Temporal Cortex | **0.57 [0.27,0.88] p=2e-04** | -0.07 [-0.33,0.18] p=0.58 | **-0.47 [-0.78,-0.15] p=3e-03** | -0.04 [-0.19,0.11] p=0.58 |
| Inferior Parietal Cortex | **0.85 [0.66,1] p=0e+00** | 0.15 [-0.16,0.46] p=0.34 | **-0.8 [-1.1,-0.45] p=8e-06** | 0.13 [-0.14,0.39] p=0.34 |
| Superior Parietal Cortex | **0.76 [0.53,0.99] p=4e-11** | -0.22 [-0.49,0.04] p=0.1 | **-0.52 [-0.83,-0.22] p=7e-04** | -0.17 [-0.38,0.04] p=0.11 |
| Amygdala | **0.49 [0.15,0.84] p=5e-03** | **-0.56 [-0.79,-0.33] p=2e-06** | -0.03 [-0.3,0.23] p=0.8 | **-0.27 [-0.5,-0.05] p=0.02** |
| Hippocampus | **0.6 [0.2,1] p=4e-03** | **-0.58 [-0.74,-0.41] p=1e-11** | **-0.46 [-0.69,-0.24] p=6e-05** | **-0.35 [-0.6,-0.09] p=8e-03** |
| Striatum | 0 [-0.31,0.32] p=0.98 | -0.07 [-0.28,0.14] p=0.5 | **-0.37 [-0.57,-0.16] p=4e-04** | 0 [-0.02,0.02] p=0.98 |

In sensitivity analyses, we assessed the role of TSPO in the context of amyloid positive ADRD and amyloid negative ADRD separately. Amyloid positive ADRD, but not amyloid negative ADRD, had a greater frequency of APOE4 compared to controls (Supplemental Table 1). Elevated amyloid and tau were the most widespread changes in amyloid positive ADRD, whereas the elevated TSPO was the most widespread change in amyloid negative ADRD (Supplemental Table 2). For TSPO and tau associations, there were positive associations that survived amyloid and neurodegeneration adjustment in amyloid positive ADRD, but similar associations did not survive adjustment in amyloid negative ADRD (Supplemental Table 3). TSPO mediated amyloid-associated tau as well as tau spreading across Braak regions in amyloid positive ADRD. TSPO also mediated tau-associated neurodegeneration in the hippocampus and amygdala in amyloid negative ADRD (Supplemental Table 4). Notably, in amyloid negative ADRD, the hippocampus had both elevated TSPO and tau. There was a negative association between TSPO and amyloid in the hippocampus for amyloid negative ADRD, driving the negative effect in the overall sample (Figure 2). This was additionally captured in the mediation analyses such that lower amyloid was associated with greater TSPO, which in turn was associated with greater tau in the hippocampus for amyloid negative ADRD. Therefore, the hippocampus was a unique region for TSPO, tau, and neurodegeneration in the absence of amyloid positivity.

## Discussion

AD-associated inflammatory alterations, measured with TSPO PET, were colocalized with tau to a greater spatial extent than amyloid and neurodegeneration, was associated with the magnitude of tau burden and neurodegeneration but not necessarily with amyloid, and was a mediator of tau-associated neurodegeneration. While typical Alzheimer’s disease is conceptualized as an amyloid-induced tauopathy, elevated TSPO was present and correlated with tau burden and neurodegeneration, particularly in the hippocampus, even in amyloid negative individuals with ADRD. Therapeutic intervention strategies should consider AD-associated inflammatory alterations as an alternative or additional target[30].

Amyloid and tau have distinct effects on microglia morphology, a proxy measure for microglia function, being ameboid near amyloid plaques and dysmorphic near tau neurofibrillary tangles[31, 32]. Cytokine-induced alterations in gene, protein, and surface receptor expression towards pro-inflammatory processes can contribute to the production, secretion, post-translation modification, aggregation, and spreading of amyloid and tau, contributing to their downstream effects on neurodegeneration and cognitive decline[1, 2]. Microglia can promote amyloid aggregation via APOE4-related pathways[33] and microglia depletion can prevent tau spreading via endosome-related pathways[34]. Further, crosstalk among dysfunctional neurons, microglia, astrocytes, and oligodendrocytes via complement cascades, inflammasome production, and other molecular components can modify these pathways[35, 36]. Additionally, build-up of insoluble lipid debris within microglia in the context of neurodegeneration can also lead to microglia dysfunction, feeding back into amyloid and tau pathology and/or neurodegeneration[37]. Rather than gaining pathology promoting functions, microglia can lose surveillance and phagocytosis function (i.e., senescence)[38], allowing amyloid and tau pathology to accumulate unchecked. The complexity of specific responses of microglia to particular targets in a given microenvironment is reflected in the nomenclature (for review, see [39]). Here, we broadly interpret TSPO PET signal to reflect AD-association inflammatory alterations that depend on the microenvironment (i.e., pathologies/co-pathologies), but is strongly linked to tau pathology and leads to neurodegeneration across ADRD diagnoses.

In this sample of older adults, there was a wide range of ER176 SUVR even in amyloid negative, tau negative controls. Hence, we chose to categorize individuals as TSPO positive if they were above the mean in controls (rather than above 1-2 standard deviations above the mean as is typical for biomarkers of AD pathology). TSPO positivity was observed in 8/10 of amyloid positive, tau positive individuals with ADRD, 2/2 amyloid negative, tau positive individuals with ADRD, and 5/5 amyloid negative, tau negative individuals with ADRD. In the context of early-onset MCI and less clinical heterogeneity (i.e., less expected copathology), TSPO PET was elevated in AD-specific regions[17, 18]. Here, amyloid negative, tau negative, TSPO positive individuals with ADRD could have unmeasured pathology (e.g., TDP-43 in LATE) or systemic risk factors that account for elevated TSPO without AD pathology. Alternatively, this could suggest that elevated TSPO can precede and promote AD pathology[10, 17]. Studies have demonstrated that AD-associated inflammatory alterations precede AD pathology in autosomal dominant AD[40], AD in adults with Down syndrome[41], and late-onset AD[42]. Longitudinal TSPO PET data capturing conversion to amyloid and/or tau PET positivity is needed.

We investigated individual AD-related regions rather than composite Braak stage ROIs to capture the spatial heterogeneity across ADRD. Our findings suggest that there may be a specific TSPO association with tau, independent of amyloid and neurodegeneration. First, we observed associations between TSPO and tau in the hippocampus, amygdala, middle inferior temporal gyrus, inferior parietal cortex, and superior parietal cortex. After accounting for amyloid and neurodegeneration, greater TSPO was still associated with greater tau burden in the inferior parietal cortex, a region that may be common to our two largest diagnostic groups—MCI/AD and PCA[43]. Second, mediation analyses demonstrated that elevated TSPO in response to elevated tau can lead to neurodegeneration in the hippocampus and amygdala, even within amyloid negative individuals with ADRD. Together, this suggests that AD-associated inflammatory alterations can be observed in relation to tau pathology without microglia simply being recruited to clear amyloid plaques and/or cellular debris (i.e., neurodegeneration). However, we cannot rule out the presence of secreted phospho-tau[32, 44], rather than tau neurofibrillary tangles measured with PET, as the driver of these TSPO-related pathways.

We also observed differential results that may reflect the complexity of AD-associated inflammatory alterations. In the cingulate gyrus, greater tau was associated with greater TSPO, which was in turn associated with greater gray matter volume, suggesting a neuroprotective effect of TSPO in the overall sample. Whether this protective effect is specific to the disease stage, brain region, or microglia subtype needs to be further explored. We found that TSPO mediated amyloid-associated tau in the middle inferior temporal gyrus, inferior parietal cortex, and superior parietal cortex, but only in the amyloid positive ADRD subgroup. Previous work suggests the link between amyloid and tau may be an astrocyte-dependent pathway[45], although astrocytes may be captured to some extent with TSPO PET. Further, we found that TSPO mediated tau spreading across Braak regions in the amyloid positive ADRD subgroup, which has been demonstrated in MCI/AD[10, 17, 34], but not in the amyloid negative ADRD subgroup, as tau spreading may follow a different spatial and temporal pattern compared to Braak staging outside of amyloid positive MCI/AD. TSPO mediated tau-associated neurodegeneration was spatially limited to the hippocampus, amygdala, and fusiform gyrus in the overall group and was driven in the hippocampus and amygdala by the amyloid negative ADRD subgroup. TSPO mediated amyloid-associated tau and TSPO mediated tau spreading may be dependent on amyloid positivity, whereas TSPO mediated tau-associated neurodegeneration may not.

Anti-amyloid treatments normalize the amyloid plaque burden[46, 47] and may prevent subsequent buildup and spreading of tau neurofibrillary tangles through TSPO-related pathways in individuals with amyloid-positive ADRD. However, the normalization of amyloid may not prevent TSPO mediated tau-associated neurodegeneration, which was present even in amyloid negative ADRD and may contribute to the pseudo-atrophy[48] thought to be related to amyloid plaque clearance from the parenchyma and/or a reduction in AD-associated inflammatory alterations. Anti-amyloids further reduce secreted phospho-tau by approximately 20%, and the contribution of soluble tau should be incorporated in future studies.

Limitations of the study include cross-sectional analysis, relatively small sample size, lack of APOE as a covariate, and lack of other measures of ADRD pathology and inflammation. While this is a cross-sectional analysis, we assessed mediations in the opposite direction to infer causality. Ongoing longitudinal data collection will allow us to additionally use temporality to infer causality. APOE genotype likely plays a critical role, along with TREM2, in microglia phagocytosis of amyloid[33]. Genetic data was not yet available for all participants and was missing more often in the ADRD group compared to the control group precluding any sensitivity analyses in those with APOE information. Sensitivity analyses suggest that APOE may be more important in amyloid positive ADRD than amyloid negative ADRD. Commonly co-occurring pathology, including cerebrovascular disease, TDP43, and α-synuclein, could be contributing to the TSPO PET signal. TSPO PET, which we broadly interpret as AD-associated inflammatory alterations, may provide transdiagnostic information through its magnitude of change and spatial distribution, similar to FDG PET for ADRD. A larger analytic sample with broad biomarker characterization is needed to provide mechanistic insight into these findings. Still, the elevated density/recruitment of microglia in AD-related brain regions, particularly the hippocampus, across different brain microenvironments provides valuable supporting information for future mechanistic investigations.

In conclusion, AD-associated inflammatory alterations, measured with TSPO PET using a simplified acquisition and quantification scheme, were present in individuals with ADRD diagnoses in a spatial pattern following tau, associated with tau burden accounting for amyloid and neurodegeneration, and mediated tau-associated neurodegeneration across various ADRD diagnoses. ‘AD-associated inflammatory alterations’ may additionally be involved in the link between amyloid and tau as well as tau spreading given the presence of elevated amyloid. Future work should focus on understanding the cellular and molecular components of these AD-associated inflammatory alterations, particularly in the hippocampus, towards a unified framework incorporating the pathogenic and immunogenic pathways in ADRD.

## Data Availability

All data produced in the present study are available upon reasonable request to the authors.

## Data Availability

All data produced in the present study are available upon reasonable request to the authors.

## Conflict of Interest Statement

WCK has a consulting agreement with Cerveau Technologies and an employee of Esai. SR is an employee of Life Molecular Imaging. However, Cerveau, Esai, and Life Molecular Imaging were not involved in the study design or interpretation of these results. WCK and SR contributed to the design, collection, and analysis of data while at Columbia University Irving Medical Center. No authors have conflicts of interest to report.

## Funding/Acknowledgements

This work was funded by NIA R01 AG063888 and R00 AG065506. Research reported in this publication was supported by the National Institute on Aging of the National Institutes of Health under Award Number P30AG066462. The content is solely the responsibility of the authors and does not necessarily represent the official views of the National Institutes of Health.

## Notes

### Clinical Trial

NCT04576793

### Funding Statement

This work was funded by NIA R01 AG063888 and R00 AG065506, and is registered on ClinicalTrials.gov under NCT04576793. Research reported in this publication was supported by the National Institute on Aging of the National Institutes of Health under Award Number P30AG066462. The content is solely the responsibility of the authors and does not necessarily represent the official views of the National Institutes of Health.

### Author Declarations

The Institutional Review Board of Columbia University Irving Medical Center gave ethical approval for this work.

