## Supplemental Materials for "Alzheimer^’^s-associated inflammatory alterations mediate tau-associated neurodegeneration in limbic and temporal regions across clinical variants of Alzheimer^’^s disease"

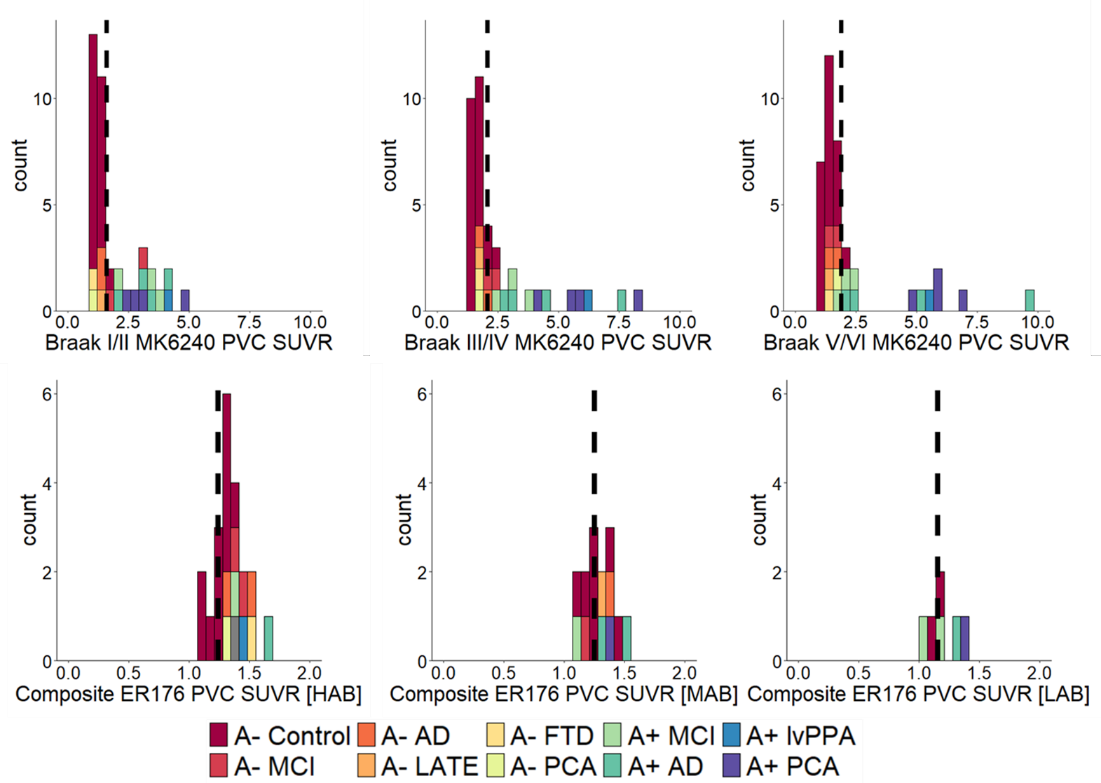

Supplemental Figure 1. Histogram of MK6240 PVC SUVR across Braak I/II, Braak III/IV, and Braak V/VI (top row) and ER176 PVC SUVR across hippocampus and a composite brain ROI (bottom row). The dashed black line indicates mean+2SD for MK6240 PVC SUVR in controls and mean for ER176 PVC SUVR in controls.

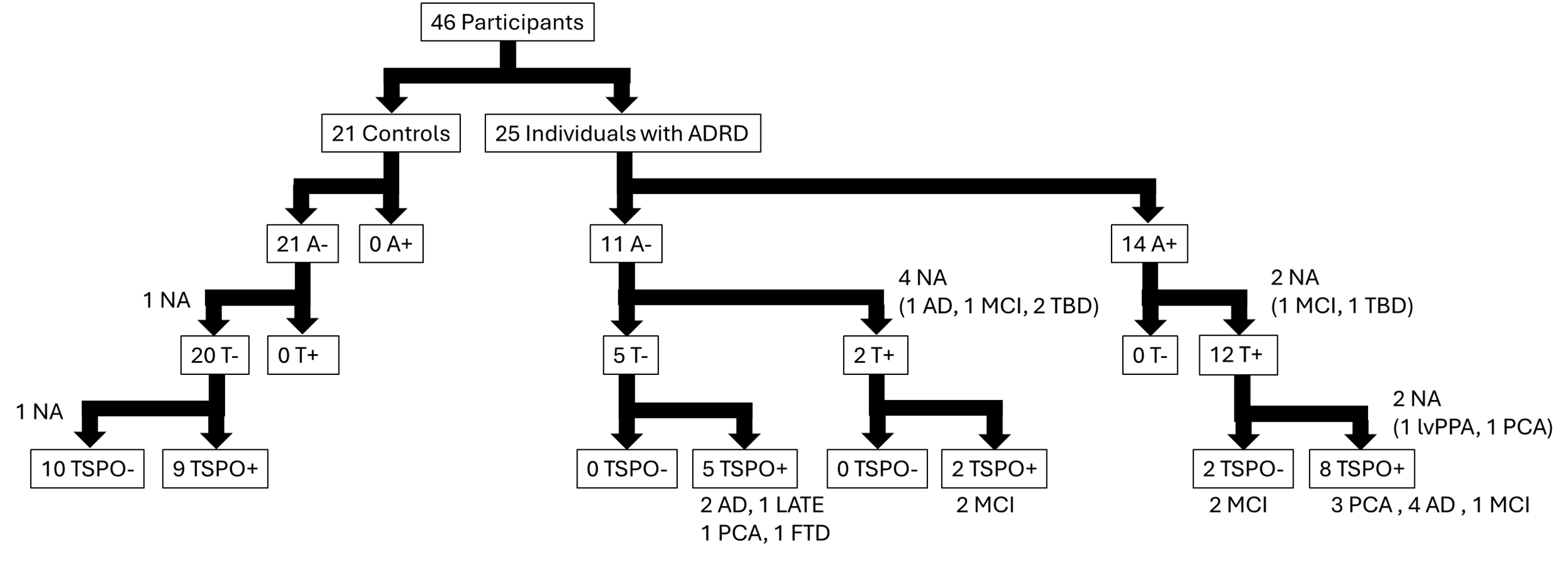

Supplemental Figure 2. Flow chart of participants by biomarker positivity and ADRD diagnosis. NA indicates individuals with partial PET acquisition (e.g., NA after amyloid positivity and before tau positivity indicates an individual had an amyloid PET, but not a tau PET yet). Multimodal PET acquisition is ongoing. TBD indicates individuals scored worse than 1.5 SD below the mean, but have not yet gone to ADRC case consensus for a specific ADRD diagnosis.

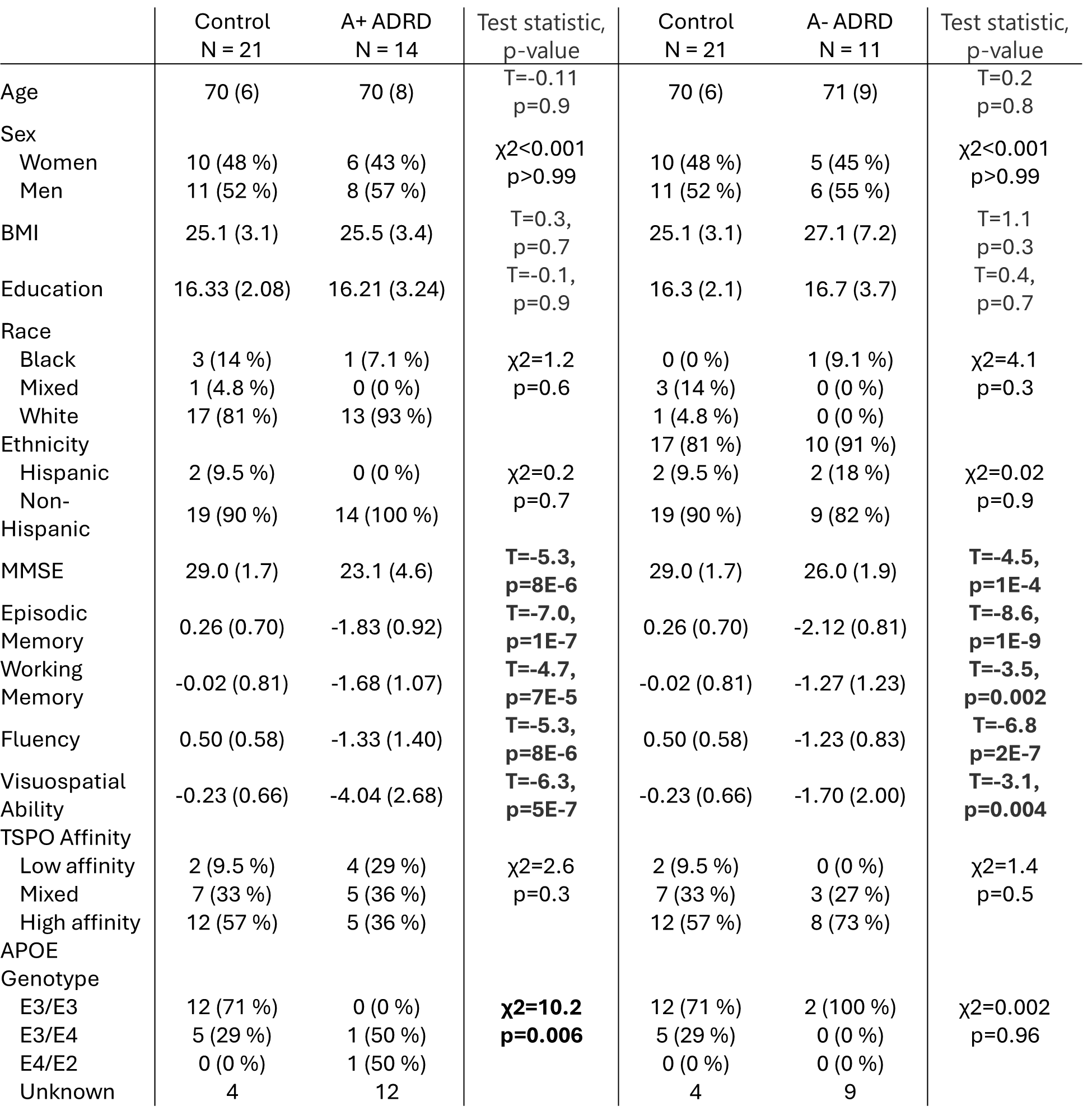

Supplemental Table 1. Demographic characteristics for controls and amyloid positive individuals with ADRD (first set of sensitivity analyses; left) and for controls and amyloid negative individuals with ADRD (second set of sensitivity analyses; right).

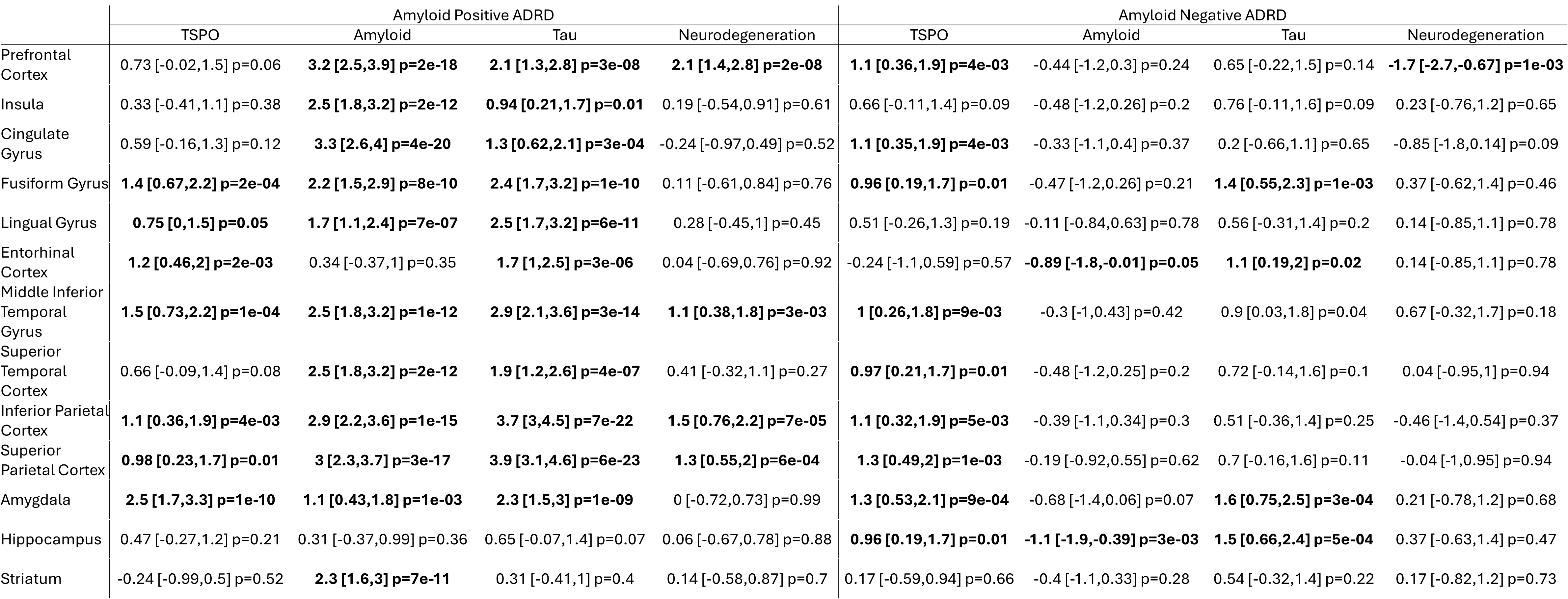

Supplemental Table 2. Standardized effect sizes between amyloid positive individuals with ADRD and controls (left) as well as amyloid negative individuals with ADRD and controls (right). Note that Neurodegeneration is inverted (Control-ADRD) relative to other biomarkers for direct comparison across standardized effect sizes.

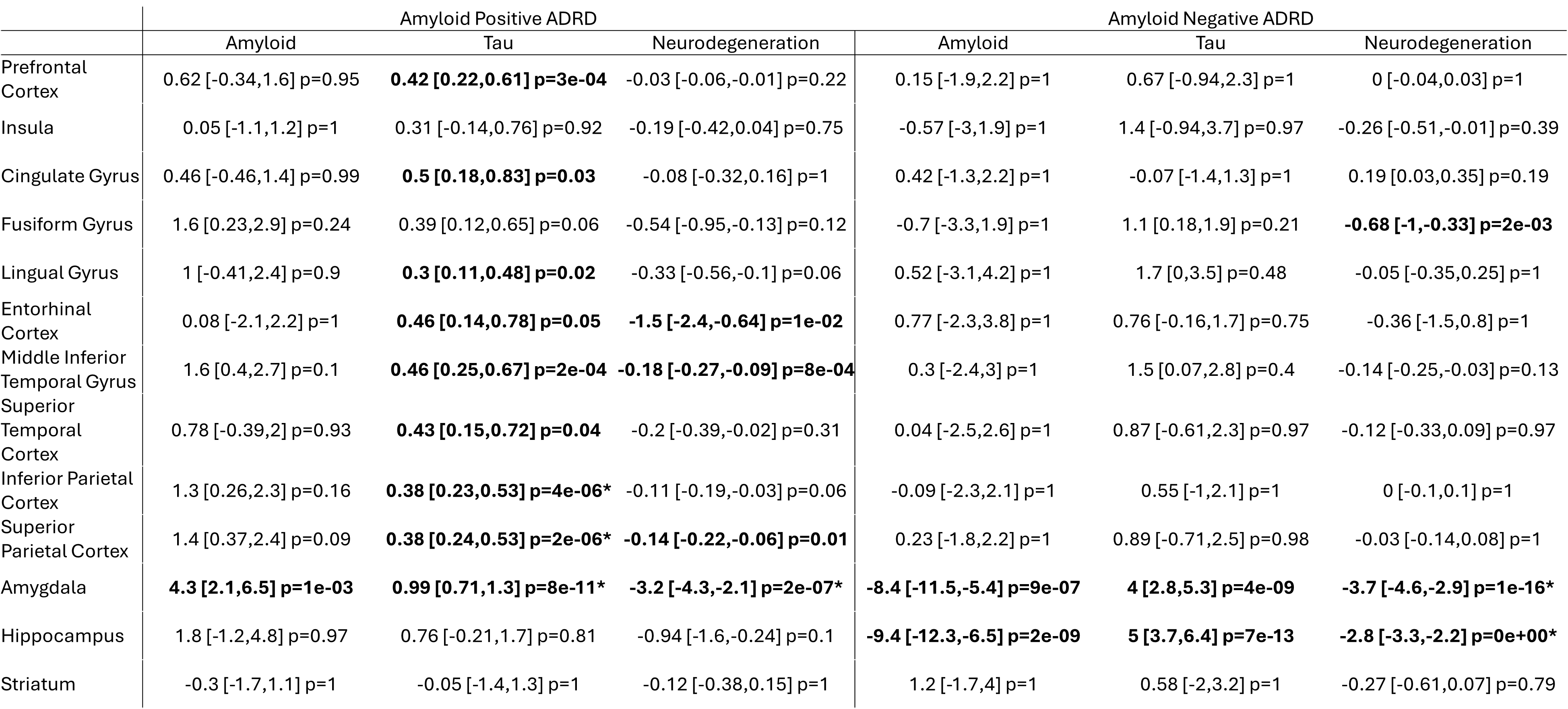

Supplemental Table 3. Standardized effect sizes for pairwise biomarker models for amyloid positive individuals with ADRD and controls (left) as well as amyloid negative individuals with ADRD and controls (right). Asterisks indicate associations that survive adjustment in the multiple biomarker model.

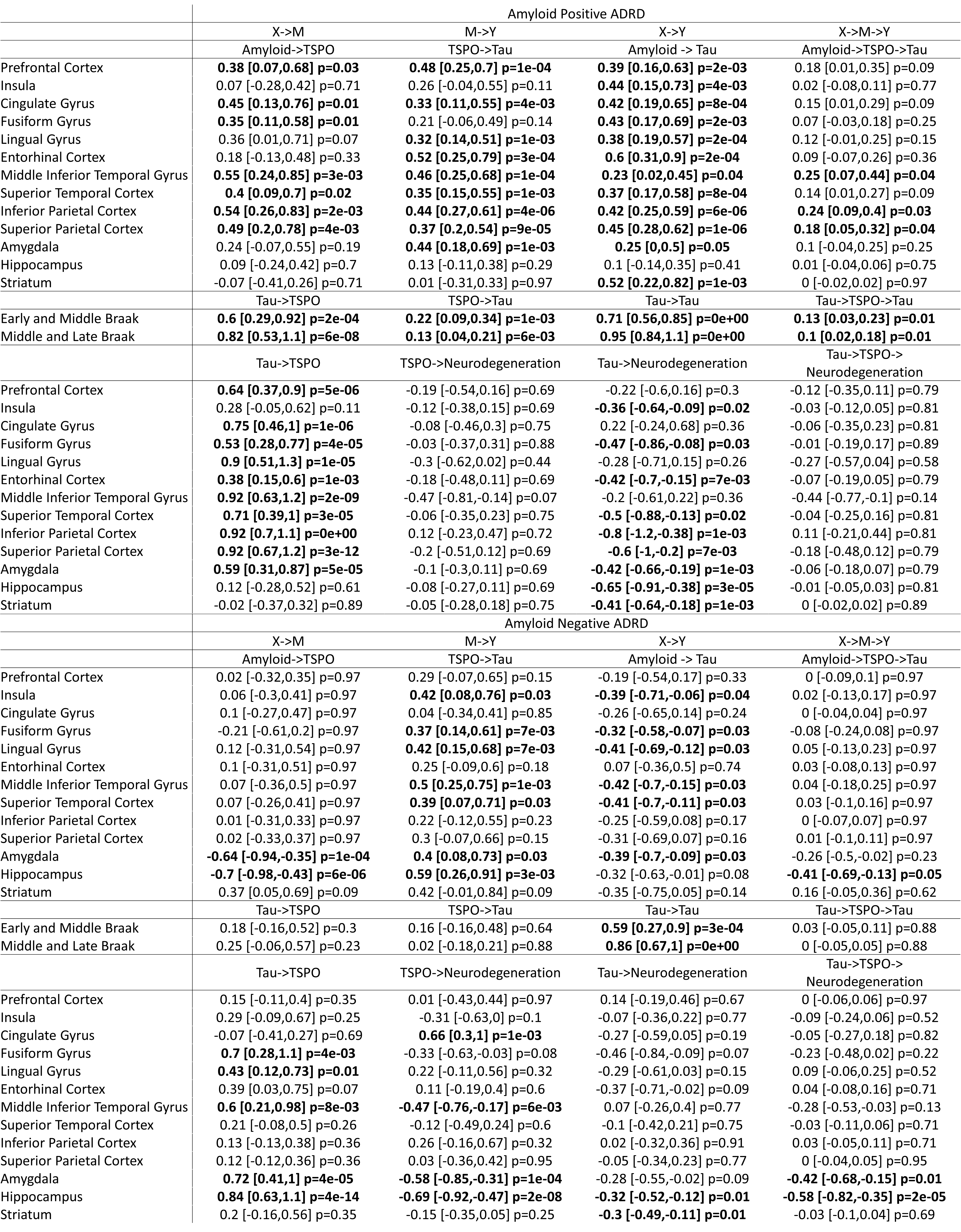

Supplemental Table 4. Standardized effect sizes for TSPO mediation models for amyloid positive individuals with ADRD and controls (top) as well as amyloid negative individuals with ADRD and controls (bottom).
